# Youth co-creation in health interventions: a systematic review and meta-analysis

**DOI:** 10.1101/2025.10.20.25338251

**Authors:** Yusha Tao, Oluwakorede J. Adedeji, Yoshiko Sakuma, Jamie Conklin, Malida Magista, Minn Thit Aung, Komang Gde Ardi Pradnya Septiawa, Esteban Ortiz Prado, Md. Kaoser Bin Siddique, Nwadiuto O. Azugo, Aishat A. Koledowo, Anita Walker, Sarah Chamouni, Juan S Izquierdo Condoy, Jorge Eduardo Vasconez Gonzales, Guillermo Jose Prieto, Isaac Alexander Suarez Sangucho, Adedayo Adeboye, Linet Mutisya, Jackie Nono, Kovey Mawuli, Ogechukwu Benedicta Aribodor, Jana Deborah Mier, Yusuf Babatunde, Chunyan Li, Ucheoma C. Nwaozuru, Day Suzanne, Eneyi E. Kpokiri, Weiming Tang, Damilola Walker, Joseph D. Tucker

## Abstract

**Introduction:** Co-creation is an iterative, bidirectional collaboration between researchers and laypeople to create knowledge. Co-creation has increasingly been recognized as an effective strategy for developing youth (10-35 years old) health interventions. This systematic review and meta-analysis examines the effectiveness of youth co-created interventions using quantitative approaches.

**Method:** We followed the Cochrane Handbook and searched PubMed, CINAHL, and Global Health on December 4, 2024, for studies reporting on youth co-creation in health. Studies were included if they involved youth co-creators in planning, designing, implementing, or evaluating interventions and reported quantitative health-related outcomes. We extracted data on youth engagement, implementation, and health outcomes, required resources, and implementation factors. Random-effects meta-analysis was used to pool the effects of the co-creation interventions on health outcomes. The study was registered in PROSPERO, CRD42024615528.

**Results:** From 17,869 citations, 112 studies were included (75,906 participants). Studies were in high-income countries (97 studies, 86.6%), middle-income countries (12 studies, 10.7%), and low-income countries (two studies, 1.8%). Interventions focused on mental health (36 studies, 32.1%), physical activity (19, 17.0%), and sexual health (11 studies, 9.8%). Most studies focused on adolescents aged 10–19 years (71 [63.4%]). Co-creation outputs included digital platforms (30.4%), toolkits or curricula (28.6%), and creative media (25.9%). The co-creation phase ranged from one-time sessions to over a year, with 43.8% of studies lasting longer than three months. Youth were universally involved as co-designers (100%) and frequently served as evaluators (40.2%), facilitators (20.5%), and presenters (13.4%), with additional roles including peer educators, outreach supporters, and co-researchers. While 63.4% of studies reported some level of youth decision-making power, only 15.2% granted lead roles and 4.5% offered final authority. Training for youth was provided in 53.6% of studies, and training for adults was provided in 38.4%, most commonly related to intervention delivery (10.7%) and facilitation or power-sharing (9.8%). However, only 49.1% of studies reported offering compensation to youth participants, and just 58.0% provided public or academic credit. Facilitators of successful co-creation included youth leadership, supportive partnerships, stakeholder engagement, flexible and relevant content, and youth training. Barriers included limited stakeholder involvement, structural barriers, insufficient resources, digital access issues, and implementation challenges. Meta-analysis showed that youth co-creation interventions improved mental health outcomes, with significant reductions in depressive (four studies, WMD: –8.63, 95% CI: –13.52 to –3.75; evidence level: low) and anxiety symptoms (four studies, WMD: –8.47, 95% CI: –12.55 to –4.38; evidence level: moderate). Co-creation interventions may increase psychological well-being (four studies; WMD: 2.31, 95% CI: –1.22 to 5.84; evidence level: very low).

**Conclusion:** Youth co-creation interventions are associated with improved health outcomes and have been implemented across diverse settings. However, youth involvement remains limited beyond design. More research is needed to optimize co-creation processes and evaluate their effectiveness.

## Introduction

Young people under 35 constitute nearly half of the world’s population,^1^ and their health is a growing global concern. They face a disproportionate burden of communicable and non-communicable diseases and injuries,^2^ while behaviors established during adolescence and early adulthood strongly influence health across the life course.^3,4^ Despite their central role, youth have historically had limited influence over the design of health research, programmes, and services that directly affect their lives.^5^ In response, co-creation is one approach that could increase youth engagement in health.^6^ Co-creation is an iterative, bidirectional collaboration between researchers and laypeople to create knowledge.

Youth co-creation offers several potential benefits. Youth co-creation can broaden youth engagement in medical research,^7,8^ tailor interventions for youth,^9^ and make policies and programs more accountable to youth.^1,10^

The World Health Organization has called for more research on participatory approaches to health innovation and evaluation, as highlighted in its 2024 Designathon Guide,^11^ and UNICEF has suggested greater inclusion of young people in health research and program development to ensure responsiveness to youth priorities.^12^ Despite these growing interests, the literature on youth co-creation in health remains fragmented. Few studies have systematically examined how co-creation has been implemented, what outcomes it has achieved, or the ethical challenges it presents. Existing reviews have largely focused on qualitative evidence and conceptual discussions, leaving quantitative assessments of intervention effectiveness largely underexplored. The absence of shared definitions and implementation standards further limits comparability across studies and hinders translation of co-creation into effective practice. Together, these calls highlight the need for a more systematic and evidence-based understanding of how youth co-creation can be effectively implemented, measured, and scaled across various contexts.

This review was organized in partnership with the TDR and UNICEF to inform the development of a practical guide on youth co-creation. The purpose of this meta-analysis and systematic review is to determine the effectiveness of youth co-created health interventions, including all studies that reported quantitative outcomes.

## Methods

This review was registered in PROSPERO (CRD42024615528). We followed the guidelines in the *Cochrane Handbook of Systematic Reviews of Interventions*^13^ and reported in accordance with the *Preferred Reporting Items for Systematic Reviews and Meta-Analyses (PRISMA)*.^14^

### Ethics

Given that this study is a systematic review and meta-analysis that synthesizes and analyzes data from previously published literature, ethical approval was not required.

### Search strategy

A medical librarian (JC) with expertise in systematic reviews assisted in developing the search strategy. We conducted a comprehensive search across three databases (PubMed, CINAHL, and Global Health) for studies reporting youth co-creation in health. Search terms included combinations of subject headings, keywords, and MeSH terms related to youths (“adolescents”, “young adults”, “youth participation”) and co-creation (“co-design”, “co-production”, “youth engagement”). There were no restrictions on the year of publication.

### Eligibility criteria

#### Youth

Youth were defined as individuals aged 10–35 years, reflecting the continuum from adolescence to early adulthood during which individuals undergo significant social, economic, and health-related transitions. Although the United Nations defines youth as 15–24 years,^15^ several regional and research frameworks have adopted broader ranges to account for extended transitions to adulthood, particularly in low- and middle-income settings. For example, the African Youth Charter defines youth as 15– 35 years.^16^ In cases where youth were a subset of a broader study population, the study or sub-analysis was included only if the intervention effectiveness was reported for a population comprising at least 75% youth participants actively engaged in co-creation activities.

#### Study design

Eligible studies included those assessing the effectiveness of co-created interventions and reporting quantitative outcomes, either alone or as part of mixed methods designs. Cluster or individually randomized trials, quasi-experimental studies, and non-controlled designs were included. There were no restrictions on disease type or geographic setting. Studies were required to involve youth as active co-creators in planning, designing, implementing, or evaluating interventions.

#### Screening and data extraction

All citations were imported into Covidence for screening, and duplicates were removed systematically.

Studies were screened in two stages. First, titles and abstracts were independently reviewed by two individuals (YT, YS, OA, KG, NA, MM, UN, AK, EO, MK, or AW), with discrepancies resolved through discussion with a third reviewer (YT, OA). Second, full-texts were independently assessed (YT and OA) to confirm eligibility. Reasons for exclusion are summarised in the Preferred Reporting Items for Systematic Reviews and Meta-Analyses (PRISMA) flow diagram (Figure 1).

**Figure 1.**
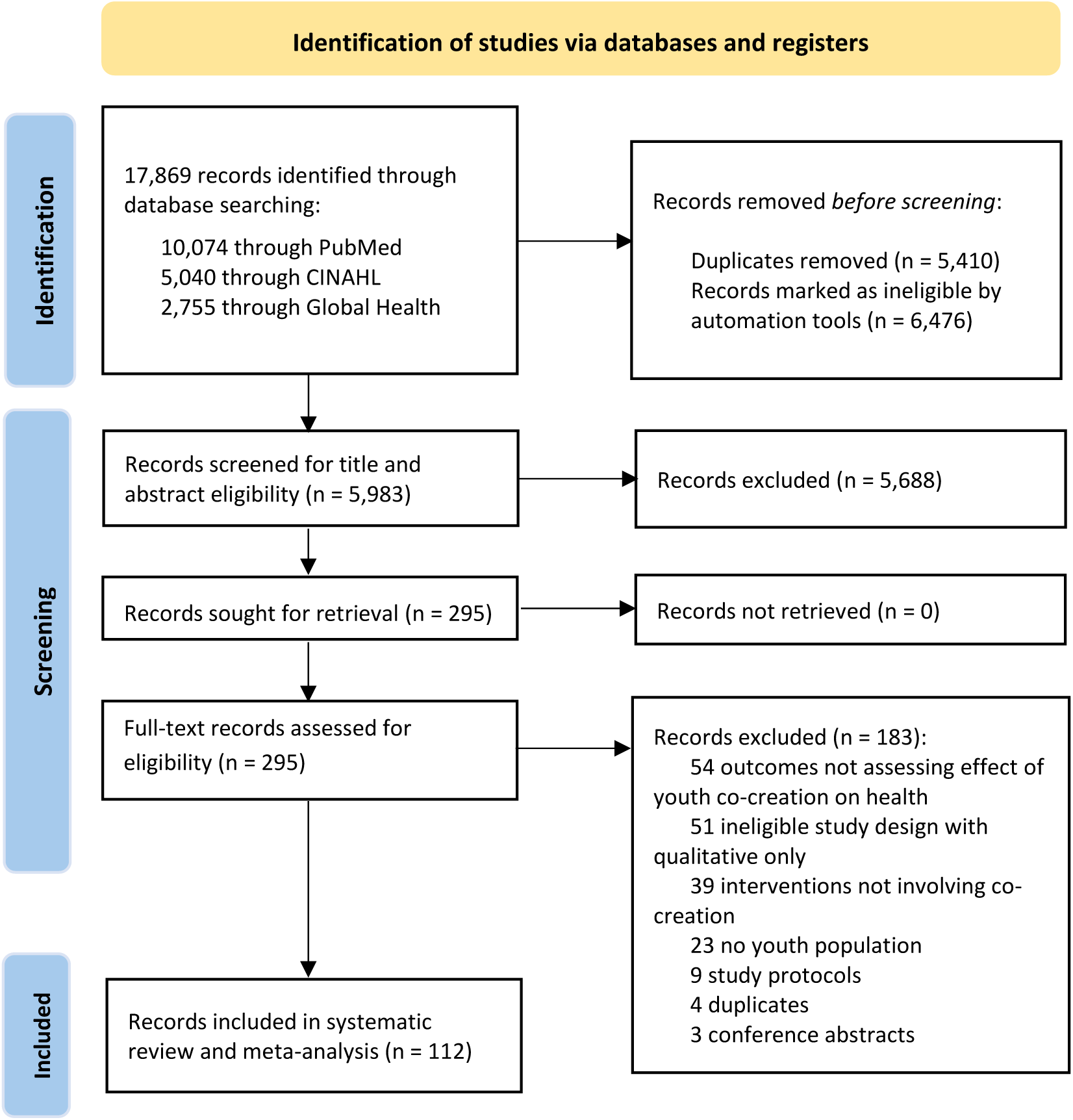
Flow diagram of study inclusion.

Data was extracted by two reviewers (YT and OA) and verified by three others (YS, MM, and MT) using a pre-tested extraction form. Extracted variables included study characteristics (country, region, setting, target population, youth age range, gender, and sample size); co-creation characteristics (youth roles, stages of engagement, decision-making power and credit, training, compensation, inclusivity, and team composition), intervention characteristics (duration, frequency, delivery mode, outputs, theoretical framework, and health domain) and outcomes (evaluation findings, acceptability, retention, resources, facilitators, barriers, strengths, and limitations).

### Analysis and evidence synthesis

Descriptive analyses were conducted to summarize study and intervention characteristics. Where studies reported comparable quantitative outcomes, random-effects meta-analysis was performed to estimate pooled effects. Weighted mean differences (WMD) with 95% confidence intervals (CIs) were calculated. Statistical heterogeneity was assessed using the I^2^ statistic, categorized as follows: low (<25%), moderate (25–75%), and high (>75%). Statistical significance was set at a two-sided p<0.05. All analyses were conducted using Stata, version 15.0 (StataCorp).

### Risk of bias and certainty of evidence

Risk of bias was assessed using design-appropriate tools: the Cochrane Risk of Bias 2 tool for randomized trials,^17^ ROBINS-I for non-randomized studies,^18^ the Johanna Briggs Institute for quasi-experimental studies,^19^ and the Mixed Methods Appraisal Tool (MMAT) for mixed-methods designs. Two reviewers (SC, EO, MM, JE, YS, IS, AA, MT, OA, JI, and JG) independently performed assessments, with disagreements resolved by consensus or through consultation with a third reviewer. The certainty of evidence was evaluated using the Grading of Recommendations Assessment, Development, and Evaluation (GRADE) framework.^20^ The certainty of study findings was rated as very low, low, moderate, or high certainty.

## Results

A total of 17,869 records were identified, of which 112 studies met the inclusion criteria (Figure 1).^21–132^ Most were conducted in high-income countries (97 [86.6%]), with a smaller proportion from low- and middle-income settings (15 [13.4%]). Study designs included quasi-experimental (43 [38.4%]), mixed-methods (42 [37.5%]), randomized controlled trials (19 [17.0%]), observational cohort or case–control (4 [3.6%]), cross-sectional (3 [2.7%]), and case series (1 [0.9%]). Populations studied included students (37 [33.0%]), youth with mental health needs (14 [12.5%]), youth with chronic physical conditions (13 [11.6%]), other or general youth populations (11 [9.8%]), racial or ethnic minority youth (10 [8.9%]), socially vulnerable youth (10 [8.9%]), health professional students (7 [6.3%]), neurodiverse youth (3 [2.7%]), pregnant or parenting youth (2 [1.8%]), youth with HIV (2 [1.8%]), mixed populations (2 [1.8%]), and youth with sexual and reproductive health needs (1 [0.9%]). Most studies focused on adolescents aged 10–19 years (71 [63.4%]), with fewer involving emerging adults aged 20–35 years (30 [26.8%]) (Table 1).

**Table 1.** Included study characteristics (n=112)

|  | Number of studies<br>(%) | Number of<br>participants |
| --- | --- | --- |
| **WHO region of study** |  |  |
| Region of the Americas | 43 (38.4%) | 9182 |
| European | 38 (33.9%) | 13454 |
| Western Pacific | 20 (17.9%) | 39770 |
| African | 5 (4.5%) | 1824 |
| South-East Asia | 4 (3.6%) | 11238 |
| Eastern Mediterranean | 1 (0.9%) | 104 |
| Multi-regions | 1 (0.9%) | 334 |
| **Country income status** |  |  |
| High income | 97 (86.6%) | 36365 |
| Upper middle income | 7 (6.3%) | 33955 |
| Lower middle income | 5 (4.5%) | 5129 |
| Low income | 2 (1.8%) | 123 |
| Mixed income | 1 (0.9%) | 334 |
| **Urban/rural** |  |  |
| Urban | 60 (53.6%) | 43123 |
| Rural | 13 (11.6%) | 5618 |
| Mixed | 18 (16.1%) | 23956 |
| Not reported | 21 (18.8%) | 3209 |
| **Study setting** |  |  |
| Educational institution | 42 (37.5%) | 59082 |
| Multi/hybrid setting | 34 (30.4%) | 8626 |
| Healthcare facility | 18 (16.1%) | 2209 |
| Community-based setting | 13 (11.6%) | 4701 |
| Digital/online setting | 5 (4.5%) | 1288 |
| **Study population** |  |  |
| General student populations | 37 (33.0%) | 59641 |
| Youth with mental health needs | 14 (12.5%) | 2712 |
| Youth with chronic physical conditions | 13 (11.6%) | 591 |
| Other/general youth populations | 11 (9.8%) | 2670 |
| Youth from racial/ethnic minorities | 10 (8.9%) | 1726 |
| Youth with social vulnerabilities | 10 (8.9%) | 3045 |
| Health professional students | 7 (6.3%) | 3088 |
| Neurodiverse youth | 3 (2.7%) | 120 |
| Pregnant or parenting youth | 2 (1.8%) | 123 |
| Youth with HIV | 2 (1.8%) | 386 |
| Mixed populations | 2 (1.8%) | 877 |
| Youth with sexual and reproductive health needs | 1 (0.9%) | 927 |
| **Age** |  |  |
| Adolescents (10-19) | 71 (63.4%) | 40382 |
| Emerging adults/youth (20-35) | 30 (26.8%) | 8668 |
| Not classified | 11 (9.8%) | 26856 |
| **Gender** |  |  |
| Mixed | 95 (84.8%) | 48660 |
| Female | 11 (9.8%) | 1655 |
| Not reported | 6 (5.4%) | 25591 |
| **Study design** |  |  |
| Quasi-experimental | 43 (38.4%) | 33410 |
| Mixed methods | 42 (37.5%) | 4757 |
| Randomized controlled trial | 19 (17.0%) | 34701 |
| Observational (cohort/case-control) | 4 (3.6%) | 2349 |
| Cross-sectional | 3 (2.7%) | 682 |
| Descriptive case-series | 1 (0.9%) | 7 |

### Stages, formats, and outputs of youth co-creation

Youth co-creation addressed a wide range of topics, most frequently mental health (36 [32.1%]), followed by physical activity, nutrition, and obesity (19 [17.0%]); chronic disease (11 [9.8%]); sexual health (11 [9.8%]); social determinants and empowerment (8 [7.1%]); substance use and addiction (7 [6.3%]); neurodevelopmental and cognitive health (5 [4.5%]); injury prevention and safety (4 [3.6%]); infectious diseases (3 [2.7%]); health services (3 [2.7%]); health education (3 [2.7%]); and environmental health (2 [1.8%]).

Youth were engaged across multiple stages of the co-creation process. Nearly all studies reported youth involvement in design (100 [100.0%]), while most involved youth in evaluation and feedback (90 [80.4%]), implementation or pilot testing of the co-creation intervention outputs (64 [57.1%]), recruitment and partnership formation (54 [48.2%]), needs assessment and planning (52 [46.4%]), dissemination and advocacy (47 [42.0%]), and training and capacity building (25 [22.3%]) (Figure 2). Youth roles ranged from advisors and team members to co-designers, facilitators, peer educators, and advocates (Figure 3).

**Figure 2.**
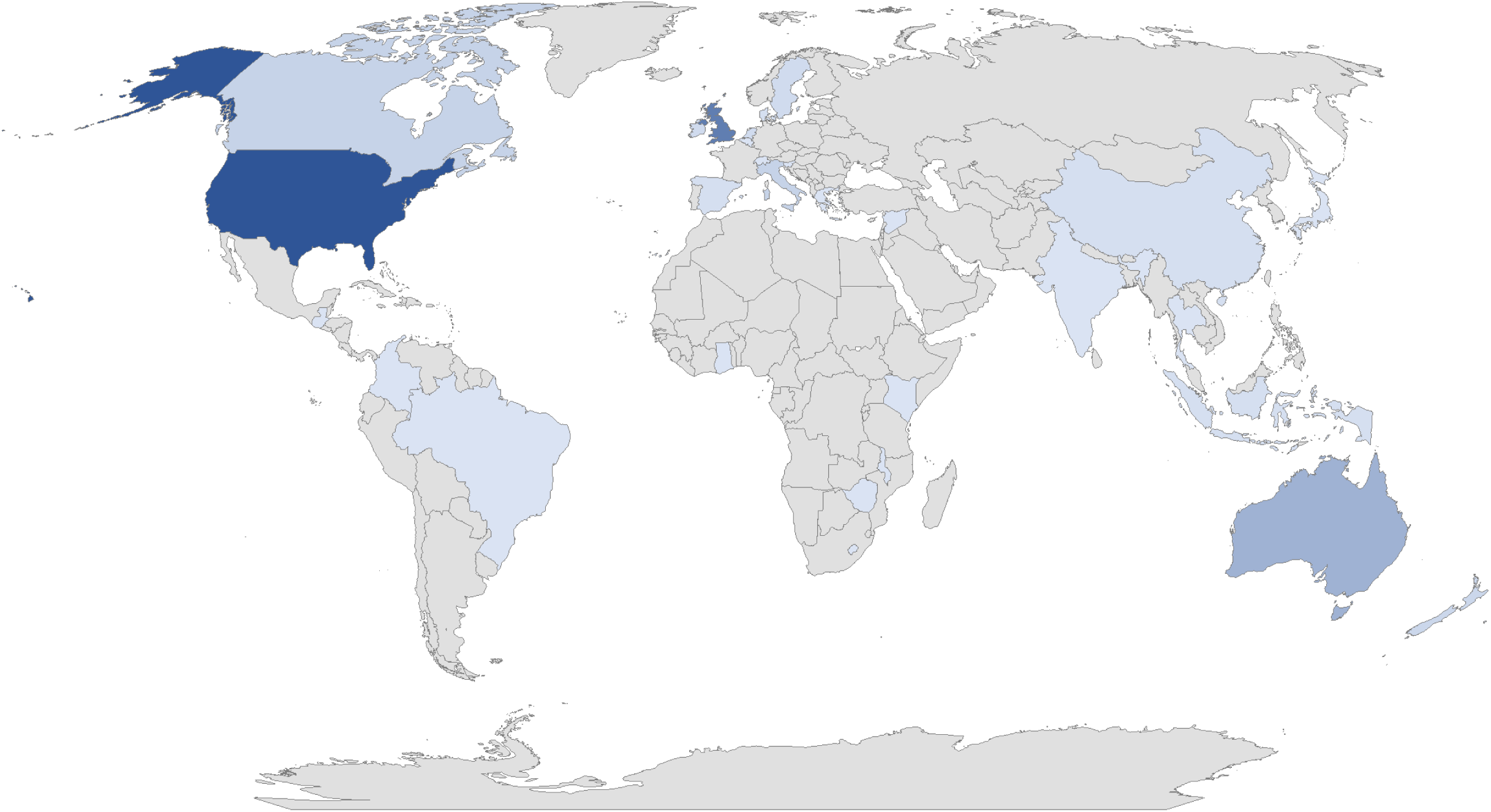
Geographical distribution of included studies (shaded in blue). WHO region: Region of the Americas – United States (n=36), Canada (n=5), Brazil (n=1), Colombia (n=1), Guatemala (n=1). **European Region** – United Kingdom (n=26), Ireland (n=3), Italy (n=5), Netherlands (n=3), Spain (n=2), Sweden (n=3), Switzerland (n=1), Belgium (n=1), Denmark ( n=2), Greece (n=1), Slovenia (n=1). **Western Pacific Region** – Australia (n=13), China (n=2), Japan (n=1), New Zealand (n=4). **South-East Asian Region** – India (n=1), Indonesia (n=2), Thailand (n=1). **Eastern Mediterranean Region** – Syrian Arab Republic (n=1). **African Region** – Ghana (n=1), Kenya (n=1), Lesotho (n=1), Malawi (n=1), Zimbabwe (n=1).

**Figure 3.**
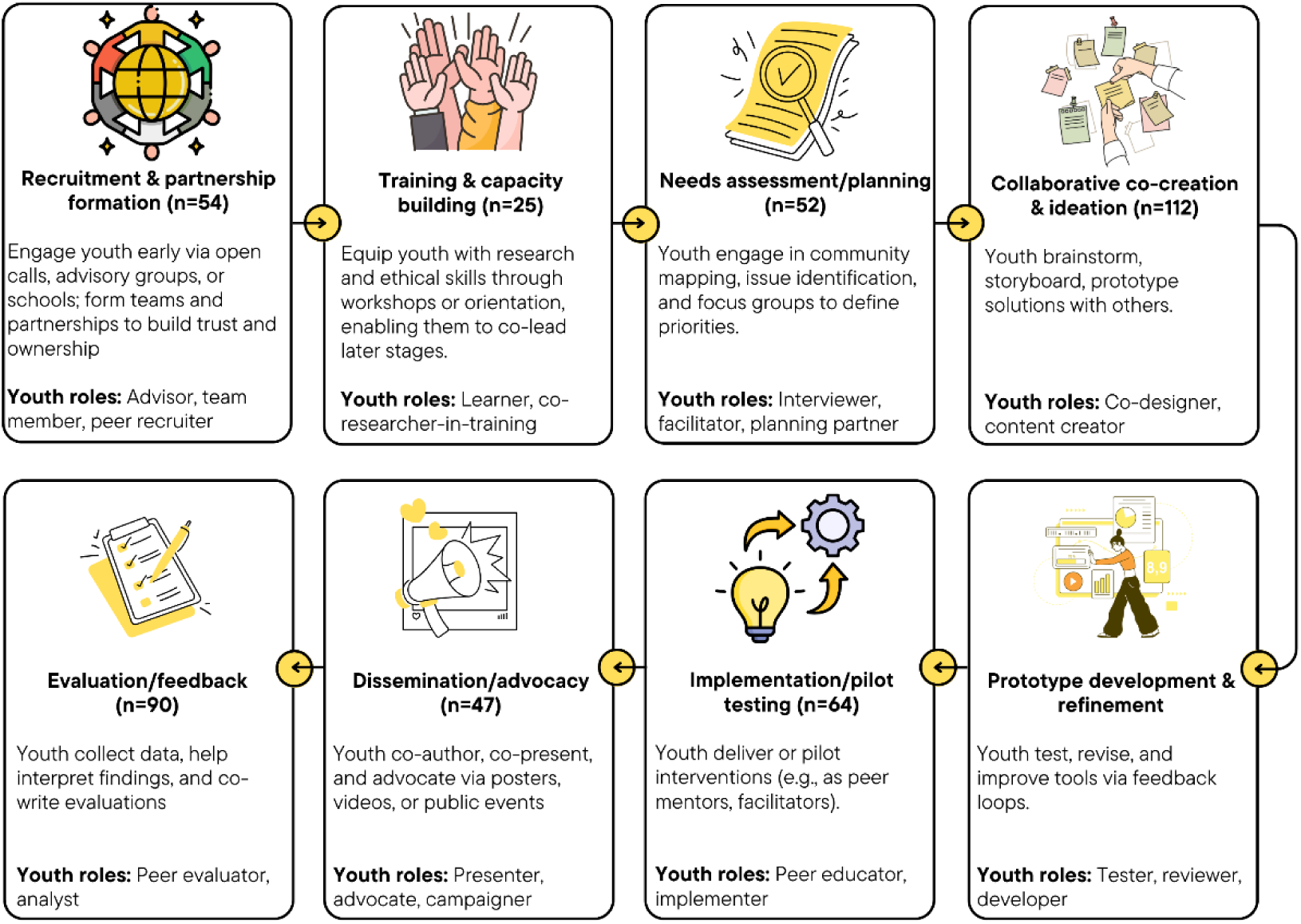
Eight-step youth co-creation framework synthesized from 112 studies This framework illustrates key stages in youth co-creation processes, derived from a synthesis of 112 included studies. n indicates how many studies reported youth participation in each step

Co-creation formats varied widely. The duration was no more than 3 months in 34 (30.4%) studies, ranging from 3 to 12 months in 32 (28.6%), and more than 1 year in 17 (15.2%). The frequency of co-creation activities ranged from weekly or more frequent sessions (38 [33.9%]) to multiple non-weekly sessions (35 [31.3%]) or one-time engagements (24 [21.4%]). Delivery modes included in-person (50 [44.6%]), online (31 [27.7%]), and hybrid (34 [30.4%]) formats.

Outputs from co-creation processes included digital platforms such as mobile phone applications or websites (34 [30.4%]); toolkits, manuals, or curricula (32 [28.6%]); creative media such as video or audio content (29 [25.9%]); campaigns or posters (22 [19.6%]); events, presentations, or workshops (18 [16.1%]); publications or reports (16 [14.3%]); games (9 [8.0%]); and policy or practice changes (4 [3.6%]) (Table 2).

**Table 2.**
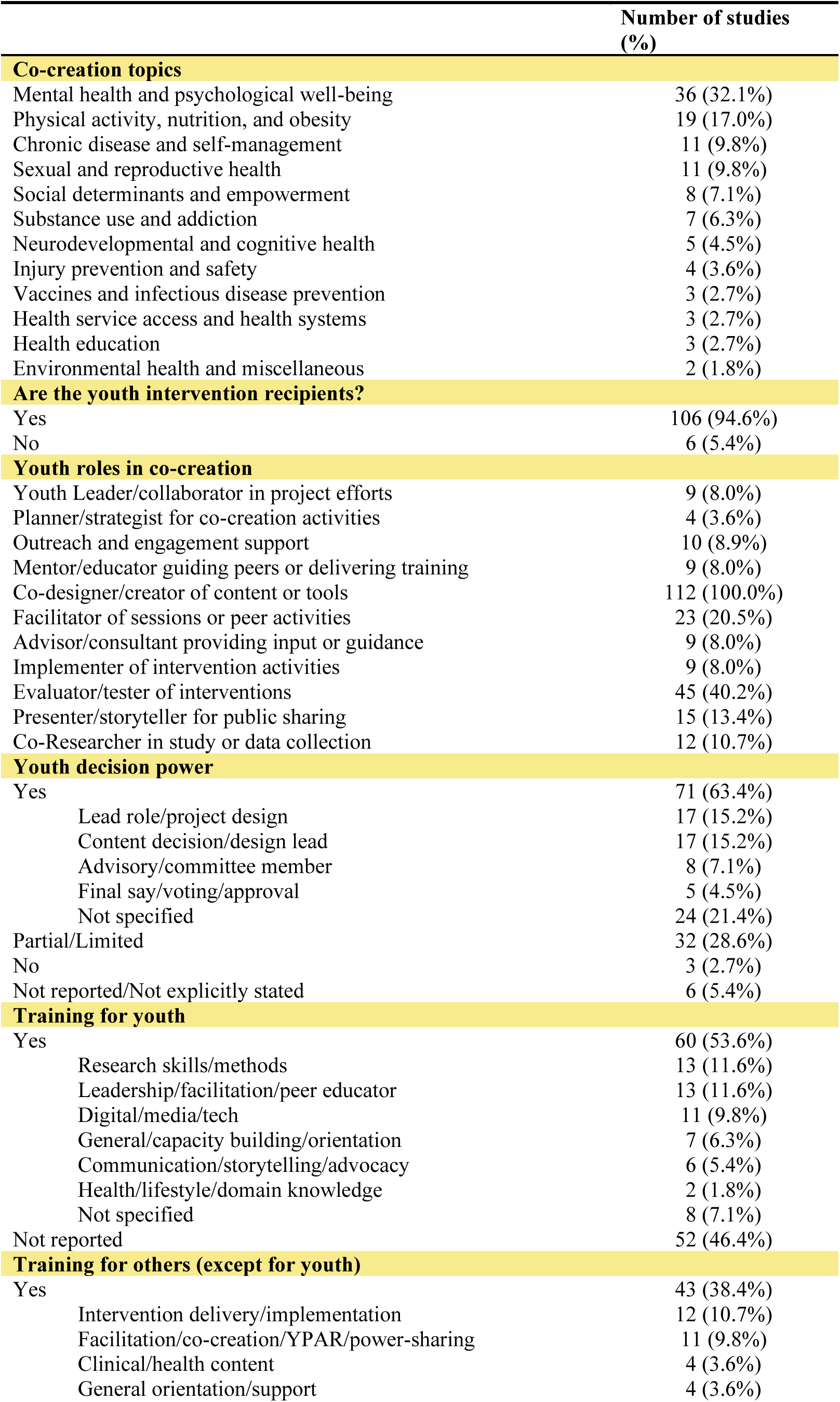
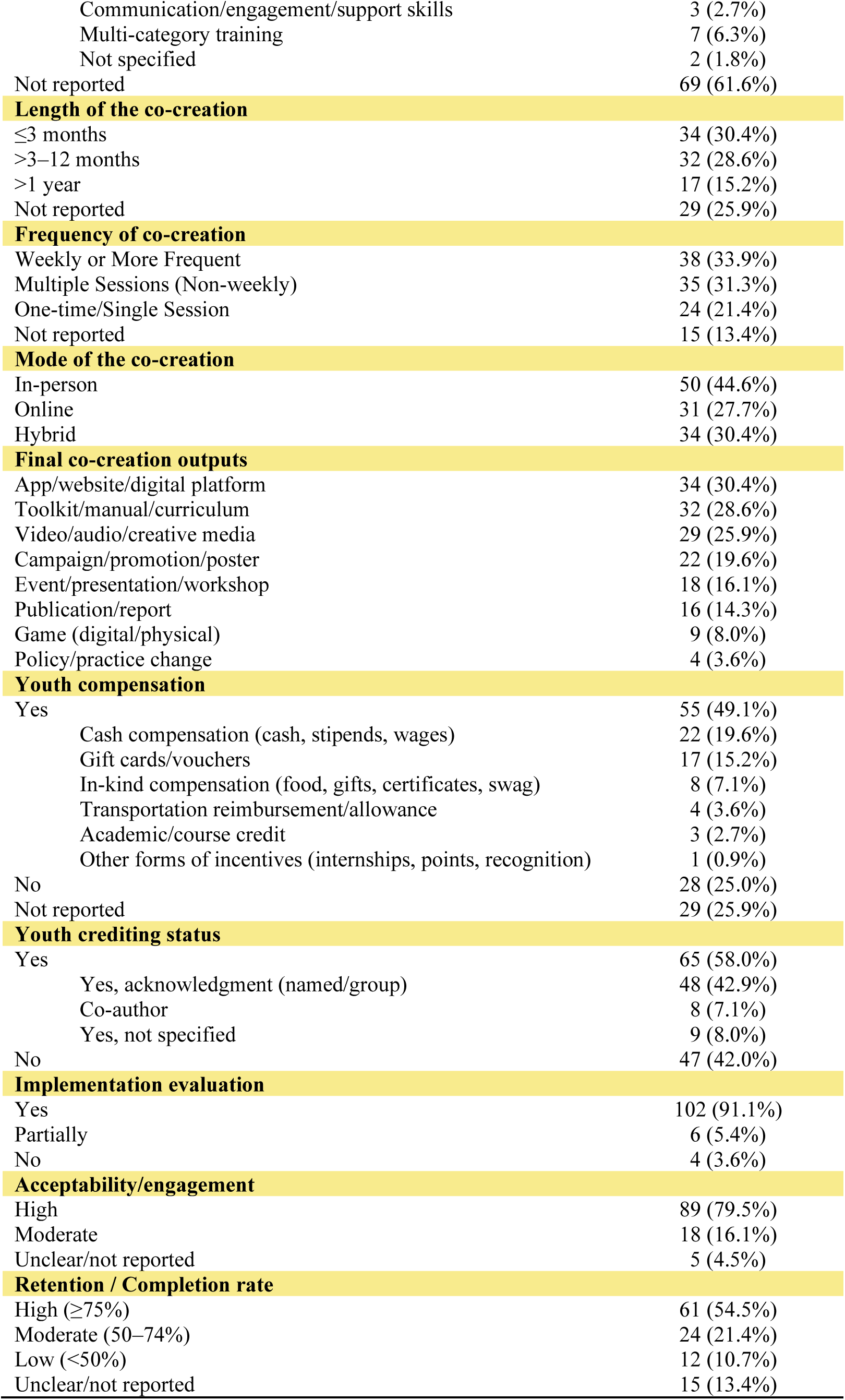
Key characteristics of youth co-creation in included studies.

|  | Number of studies (%) |
| --- | --- |
| **Co-creation topics** |  |
| Mental health and psychological well-being | 36 (32.1%) |
| Physical activity, nutrition, and obesity | 19 (17.0%) |
| Chronic disease and self-management | 11 (9.8%) |
| Sexual and reproductive health | 11 (9.8%) |
| Social determinants and empowerment | 8 (7.1%) |
| Substance use and addiction | 7 (6.3%) |
| Neurodevelopmental and cognitive health | 5 (4.5%) |
| Injury prevention and safety | 4 (3.6%) |
| Vaccines and infectious disease prevention | 3 (2.7%) |
| Health service access and health systems | 3 (2.7%) |
| Health education | 3 (2.7%) |
| Environmental health and miscellaneous | 2 (1.8%) |
| **Are the youth intervention recipients?** |  |
| Yes | 106 (94.6%) |
| No | 6 (5.4%) |
| **Youth roles in co-creation** |  |
| Youth Leader/collaborator in project efforts | 9 (8.0%) |
| Planner/strategist for co-creation activities | 4 (3.6%) |
| Outreach and engagement support | 10 (8.9%) |
| Mentor/educator guiding peers or delivering training | 9 (8.0%) |
| Co-designer/creator of content or tools | 112 (100.0%) |
| Facilitator of sessions or peer activities | 23 (20.5%) |
| Advisor/consultant providing input or guidance | 9 (8.0%) |
| Implementer of intervention activities | 9 (8.0%) |
| Evaluator/tester of interventions | 45 (40.2%) |
| Presenter/storyteller for public sharing | 15 (13.4%) |
| Co-Researcher in study or data collection | 12 (10.7%) |
| **Youth decision power** |  |
| Yes | 71 (63.4%) |
| Lead role/project design | 17 (15.2%) |
| Content decision/design lead | 17 (15.2%) |
| Advisory/committee member | 8 (7.1%) |
| Final say/voting/approval | 5 (4.5%) |
| Not specified | 24 (21.4%) |
| Partial/Limited | 32 (28.6%) |
| No | 3 (2.7%) |
| Not reported/Not explicitly stated | 6 (5.4%) |
| **Training for youth** |  |
| Yes | 60 (53.6%) |
| Research skills/methods | 13 (11.6%) |
| Leadership/facilitation/peer educator | 13 (11.6%) |
| Digital/media/tech | 11 (9.8%) |
| General/capacity building/orientation | 7 (6.3%) |
| Communication/storytelling/advocacy | 6 (5.4%) |
| Health/lifestyle/domain knowledge | 2 (1.8%) |
| Not specified | 8 (7.1%) |
| Not reported | 52 (46.4%) |
| **Training for others (except for youth)** |  |
| Yes | 43 (38.4%) |
| Intervention delivery/implementation | 12 (10.7%) |
| Facilitation/co-creation/YPAR/power-sharing | 11 (9.8%) |
| Clinical/health content | 4 (3.6%) |
| General orientation/support | 4 (3.6%) |
| Communication/engagement/support skills | 3 (2.7%) |
| Multi-category training | 7 (6.3%) |
| Not specified | 2 (1.8%) |
| Not reported | 69 (61.6%) |
| **Length of the co-creation** |  |
| ≤3 months | 34 (30.4%) |
| >3–12 months | 32 (28.6%) |
| >1 year | 17 (15.2%) |
| Not reported | 29 (25.9%) |
| **Frequency of co-creation** |  |
| Weekly or More Frequent | 38 (33.9%) |
| Multiple Sessions (Non-weekly) | 35 (31.3%) |
| One-time/Single Session | 24 (21.4%) |
| Not reported | 15 (13.4%) |
| **Mode of the co-creation** |  |
| In-person | 50 (44.6%) |
| Online | 31 (27.7%) |
| Hybrid | 34 (30.4%) |
| **Final co-creation outputs** |  |
| App/website/digital platform | 34 (30.4%) |
| Toolkit/manual/curriculum | 32 (28.6%) |
| Video/audio/creative media | 29 (25.9%) |
| Campaign/promotion/poster | 22 (19.6%) |
| Event/presentation/workshop | 18 (16.1%) |
| Publication/report | 16 (14.3%) |
| Game (digital/physical) | 9 (8.0%) |
| Policy/practice change | 4 (3.6%) |
| **Youth compensation** |  |
| Yes | 55 (49.1%) |
| Cash compensation (cash, stipends, wages) | 22 (19.6%) |
| Gift cards/vouchers | 17 (15.2%) |
| In-kind compensation (food, gifts, certificates, swag) | 8 (7.1%) |
| Transportation reimbursement/allowance | 4 (3.6%) |
| Academic/course credit | 3 (2.7%) |
| Other forms of incentives (internships, points, recognition) | 1 (0.9%) |
| No | 28 (25.0%) |
| Not reported | 29 (25.9%) |
| **Youth crediting status** |  |
| Yes | 65 (58.0%) |
| Yes, acknowledgment (named/group) | 48 (42.9%) |
| Co-author | 8 (7.1%) |
| Yes, not specified | 9 (8.0%) |
| No | 47 (42.0%) |
| **Implementation evaluation** |  |
| Yes | 102 (91.1%) |
| Partially | 6 (5.4%) |
| No | 4 (3.6%) |
| **Acceptability/engagement** |  |
| High | 89 (79.5%) |
| Moderate | 18 (16.1%) |
| Unclear/not reported | 5 (4.5%) |
| **Retention / Completion rate** |  |
| High (≥75%) | 61 (54.5%) |
| Moderate (50–74%) | 24 (21.4%) |
| Low (<50%) | 12 (10.7%) |
| Unclear/not reported | 15 (13.4%) |

### Training, compensation, credit, and decision-making power

Training for youth was reported in 60 [53.6%] studies, focusing on research skills and methods (13 [11.6%]), leadership, facilitation, and peer education (13 [11.6%]), digital and media/technology skills (11 [9.8%]), general capacity building and orientation (7 [6.3%]), communication, storytelling, and advocacy (6 [5.4%]), and health-specific knowledge (2 [1.8%]). Training for adults was reported in 43 (38.4%) studies, most focused on intervention delivery or implementation (12 [10.7%]), facilitation, or power-sharing (11 [9.8%]), clinical or health content (4 [3.6%]), general orientation or support (4 [3.6%]), and communication or engagement skills (3 [2.7%]); seven (6.3%) reported multi-category training.

Compensation for youth co-creation was described in 55 (49.1%) studies, most commonly in the form of cash stipends or wages (22 [19.6%]) and gift cards or vouchers (17 [15.2%]). Other incentives included in-kind support such as food, gifts, or certificates (8 [7.1%]), transport reimbursement (4 [3.6%]), academic or course credit (3 [2.7%]), and internships or recognition (1 [0.9%]). Youth were credited in 65 (58.0%) studies, either through named or group acknowledgment (48 [42.9%]), or co-authorship (8 [7.1%]).

Decision-making power was shown in 71 (63.4%) studies. Youth most often held leadership roles in project design (17 [15.2%]) or content decisions (17 [15.2%]), served on advisory or steering committees (8 [7.1%]), or had final voting or approval authority (5 [4.5%]) (Table 2).

### Implementation, acceptability, and retention

Most studies (102 [91.1%]) evaluated the implementation of co-created outputs. Acceptability and engagement were high in 89 (79.5%) studies and moderate in 18 (16.1%). Retention or completion rates of the co-creation were high (≥75%) in 61 (54.5%) studies, moderate (50–74%) in 24 (21.4%), and low (<50%) in 12 (10.7%) (Table 2).

### Effects of youth co-creation on mental health outcomes

Four studies assessed the effect of youth co-creation on depressive symptoms using the *Mood and Feelings Questionnaire* and were pooled in a random-effects meta-analysis (Figure 4a). Youth co-created interventions were associated with a significant reduction in depressive symptoms compared with non-co-created interventions (weighted mean difference [WMD], –8.63 [95% CI, –13.52 to –3.75]; I² = 66.1%, p = 0.032).

**Figure 4.**
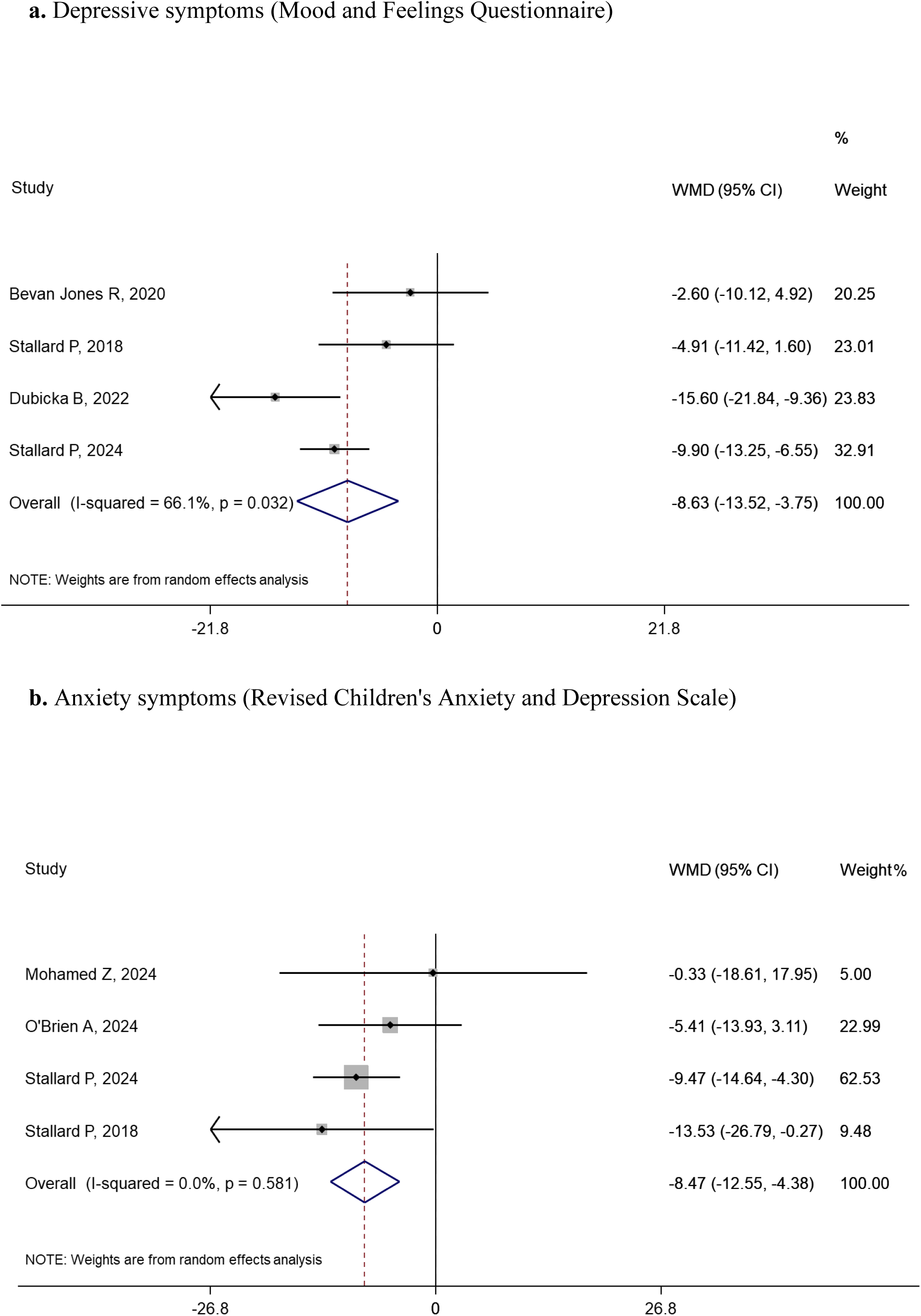
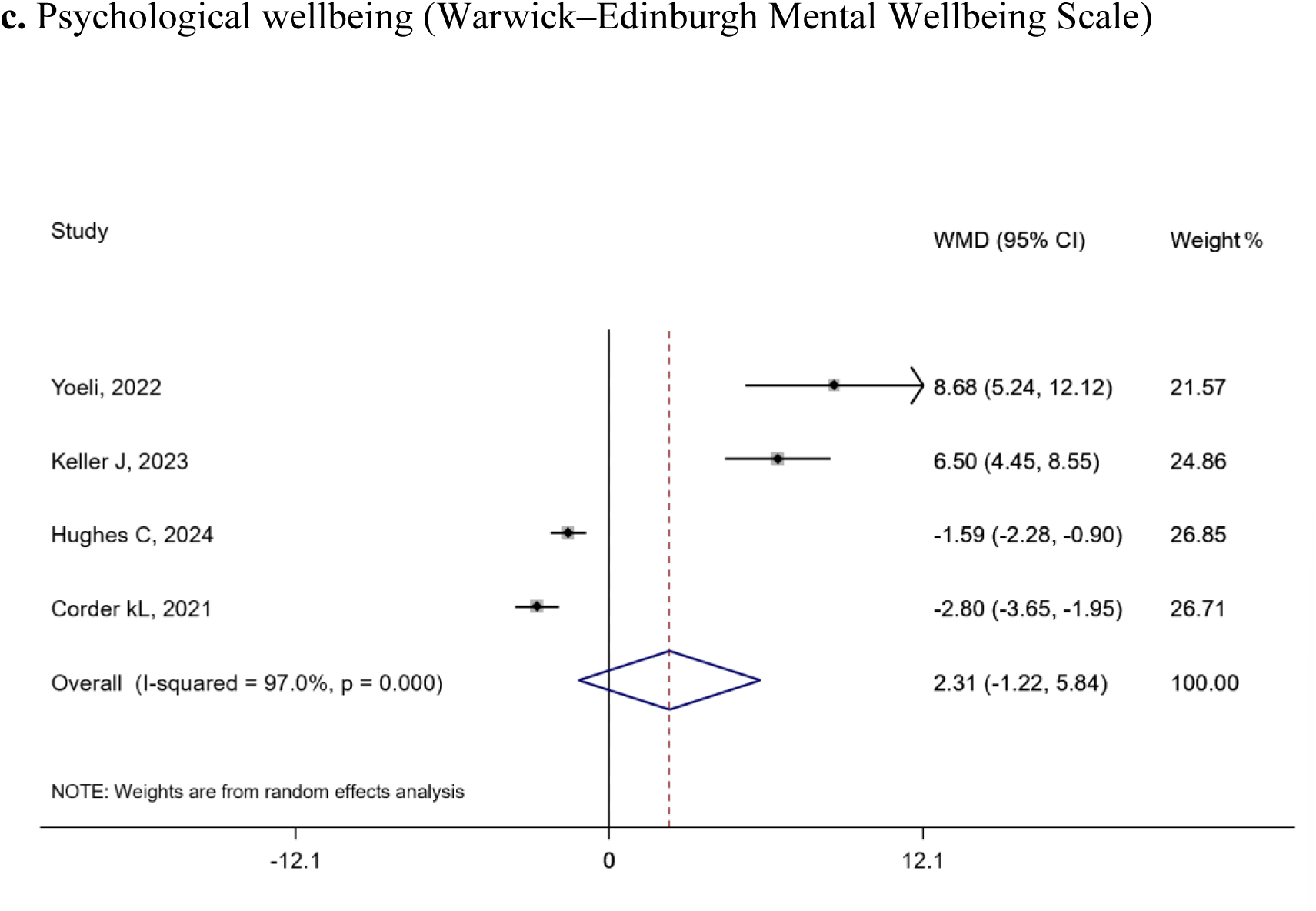
Meta-analysis of youth co-creation versus non-co-creation interventions: weighted mean difference (WMD) for (a) depressive symptoms (Mood and Feelings Questionnaire), (b) anxiety symptoms (Revised Children’s Anxiety and Depression Scale), and (c) psychological wellbeing (Warwick–Edinburgh Mental Wellbeing Scale).

Four studies evaluated anxiety symptoms using the *Revised Children’s Anxiety and Depression Scale*. Youth co-creation interventions were associated with lower anxiety symptoms (WMD, –8.47 [95% CI, –12.55 to –4.38]; I² = 0.0%, p = 0.581) (Figure 4b).

There were also four studies that assessed psychological wellbeing using the *Warwick–Edinburgh Mental Wellbeing Scale*. There was no difference in well-being observed (WMD, 2.31 [95% CI, –1.22 to 5.84]; I² = 97.0%, p < 0.001) (Figure 4c).

### Theories and frameworks guiding youth co-creation

Youth co-creation interventions were developed based on diverse theoretical frameworks (Table 3). Most were grounded in empowerment and participatory principles, including youth empowerment theory, sociopolitical development theory, youth participatory action research, and community-based participatory research. Co-design and human-centered design approaches were also widely used. Behavioral and learning theories, such as self-determination theory, social cognitive theory, positive youth development, and youth–adult partnership models, informed several studies. Additional frameworks included design thinking, gamification, realist evaluation, complexity theory, and ecological models.

**Table 3.**
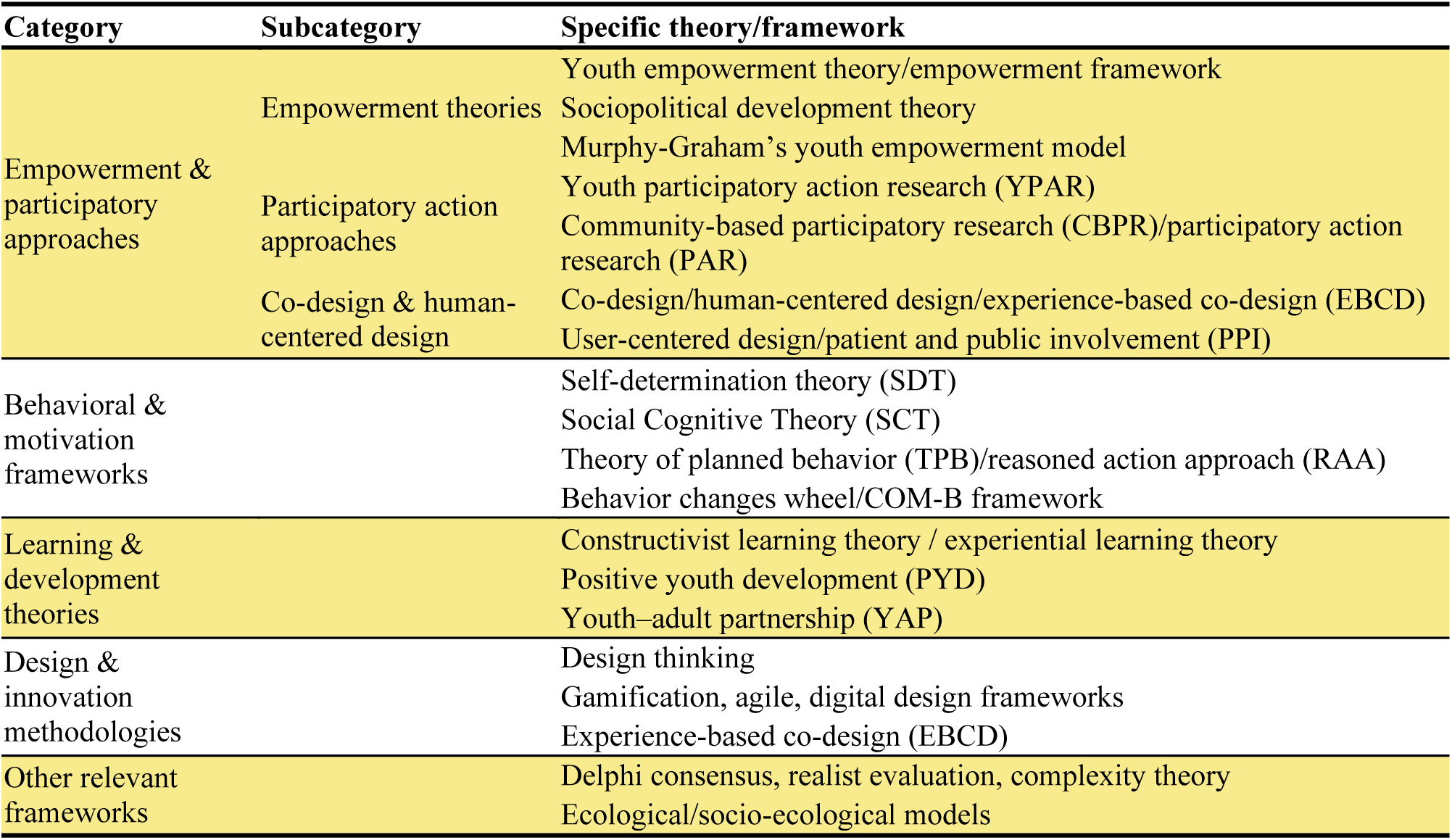
Theories and frameworks used to guide youth co-creation design。.

### Facilitators and barriers for organizing youth co-creation

Several facilitators were reported across studies. Strong youth engagement and leadership were important to ensuring meaningful youth participation (68 studies). Supportive partnerships that enabled power-sharing further strengthened co-creation processes (54 studies), while multi-stakeholder collaboration created broader support (46 studies). Relevant, flexible, and appealing content or formats helped sustain youth interest (39 studies), and training, capacity building, and supervision were essential in equipping young people to contribute effectively (31 studies).

By contrast, several barriers limited successful implementation. The most frequent challenges included limited stakeholder and participant engagement (53 studies) and resource constraints related to sample size, time, and funding (49 studies). Structural and administrative barriers were also common (38 studies), alongside technological and digital barriers that restricted participation in some contexts (33 studies). Additional difficulties, including program design and implementation challenges, hindered the consistency and scalability of co-creation approaches (27 studies).

### Strengths and limitations of using a youth co-creation

Studies highlighted several strengths of youth co-creation. Several studies discussed its capacity to empower young people to lead, make decisions, and directly influence interventions (64 studies). Co-creation increased engagement, satisfaction, and sustained participation (58 studies) and fostered participatory, peer-led, and collaborative processes in program development (50 studies). Tailoring interventions to local culture, context, and real-world settings enhanced relevance and uptake (44 studies), while promoting diversity, equity, and inclusion ensured that a broad range of youth voices were represented (36 studies).

Several limitations of using youth co-creation were also discussed. Lack of standardization across studies produced variability in practice (48 studies), and the sustainability and scalability of youth co-creation models remained uncertain (41 studies). In addition, many studies face challenges in maintaining meaningful and consistent youth engagement throughout the process (35 studies).

## Discussion

This systematic review and meta-analysis shows that youth co-creation is a promising and adaptable approach that strengthens the design and delivery of health interventions. Across studies, co-creation has enhanced innovation and relevance by integrating youth perspectives with multidisciplinary expertise, particularly in the field of mental health. It demonstrated feasibility and acceptability across a range of populations and formats, including those involving marginalized youth, suggesting its scalability within health systems. Despite diverse implementation models, shared principles of partnership, power-sharing, and contextual adaptation emerged as central to successful practice. These findings indicate that youth co-creation offers a practical framework for developing more participatory, equitable, and responsive health interventions.

Our review identified evidence that youth co-created interventions were associated with reductions in depressive and anxiety symptoms. These findings are consistent with a recent review in which 17% of youth co-design studies reported measurable improvements in youth mental health outcomes.^133^ Co-creation processes may contribute to these effects by enhancing psychological agency, strengthening social connectedness, and increasing the contextual relevance of interventions to promote mental well-being. Youth co-creation also ensured that youth and other stakeholders’ expertise were heard and valued, leading to mutual benefits and improved mental health outcomes.^133–135^

However, adequate training for both youth and adult collaborators with the skills and awareness is essential. Previous studies demonstrated that moving from consultants to co-researchers requires structured support in research literacy, communication, and collaboration skills, and a clear definition of roles and responsibilities. Explicit, accessible training improves youths’ knowledge, confidence, and readiness to engage as research partners while reducing barriers to their involvement.^136^ Training for adults is also important, for example, in mental health contexts, training in trauma-informed practices and cultural competence should be prioritized to ensure that co-creation processes are safe, inclusive, and meaningful.^137^

Evidence from our review suggests the need for embedding youth co-creation within routine governance and decision-making, rather than relying on one-off consultations. In policy and practice, meaningful youth involvement can improve relevance, equity, and health outcomes. To realize sustained benefits, co-creation should be formalized through clearly defined decision rights, dedicated resources, and organization-wide standards for reporting and accountability. We therefore advocate policy changes that provide opportunities for youth participation within the decision-making arms of governments, communities, and organizations and that invest in intergenerational partnerships to generate more inclusive, durable solutions.

This study expands the literature by systematically examining youth-led co-creation in health research across multiple domains and settings. A key strength of this review is that it maps how co-creation has been conceptualized, implemented, and evaluated, and integrates conceptual, methodological, and practical dimensions to inform future applications. By cataloging youth roles, training, compensation, credit, and decision-making power, our review provides a framework to strengthen implementation quality, equity, and accountability in youth co-creation.

We acknowledge several limitations in this review that have to be noted. First, we excluded studies that reported only qualitative findings, which may limit the comprehensiveness of our data synthesis on the topic. Second, the included studies reported a wide range of outcomes, resulting in substantial heterogeneity in the data and preventing a meta-analysis from being conducted. Third, most of the studies included in this review were conducted in high-income countries. While this may be partly attributable to our focus on studies that reported quantitative data, it highlights the need for further research in low- and middle-income countries and in diverse contexts to better understand how youth-led co-creation is conceptualized and implemented in the global settings.

In summary, this review shows that youth co-creation is a flexible and adaptable approach applied across diverse populations, health domains, and settings. It fostered innovation by engaging youth alongside interdisciplinary partners throughout intervention design and evaluation, and demonstrated feasibility, acceptability, and potential to improve outcomes. However, limited decision-making power and fair compensation for youth were often lacking. Future research should address these gaps by developing shared standards and implementation strategies that strengthen equity and impact in youth co-creation for health.

## Data Availability

All data produced in the present study are available upon reasonable request to the authors

